# Cost of Goods Sold Analysis for Manufacturing mRNA-Based Cell and Gene Therapies

**DOI:** 10.64898/2026.05.04.26352406

**Authors:** Robert D. Lieberthal, Peter Buontempo, Brittani Harmon, Anthony Omosule, Mike Washabaugh, Alex Whittaker

## Abstract

**Background:** Cell and gene therapies (CGT) represent a transformative class of medical interventions, yet their high production costs limit patient access. Understanding the structure of manufacturing costs is essential for informing policies that can expand access to these therapies.

**Objective:** This study develops and applies a cost-of-goods-sold (COGS) model to analyze the contributors to manufacturing costs for mRNA-based CGT, with application to a wide range of current and future therapies.

**Methods:** An Excel-based COGS model was constructed based on cost categories for CGT. Two mRNA-based products at commercial scale were used to populate the model: an mRNA vaccine and a therapeutic mRNA gene therapy. Cost inputs were drawn from vendor pricing, peer-reviewed and grey literature, and expert consultation with CGT manufacturing specialists. Three scenarios (worst, base, and best case) were modeled across six cost categories: materials, consumables, capital, labor, licenses, and royalties. A tornado diagram sensitivity analysis was conducted to identify key cost drivers. The mRNA vaccine was used to build and validate the model strucutre using publicly available data sources. The therapeutic mRNA therapy was used as the main use case for illustration and sensitivity analysis.

**Results:** Under base-case assumptions, the estimated cost per dose for the ther-apeutic mRNA product is $56.09, ranging from $3.68 (best case) to $383.22 (worst case). Licensing and royalty fees together account for approximately 83% of total base-case COGS ($6,996,000 and $6,960,000 per production run, respectively, out of $16,825,597 total). Excluding these fees, material costs represent the largest remaining share (61%), followed by consumables (34%), capital (4%), and labor (1%). Sensitivity analysis confirms that licensing and royalty assumptions are the dominant source of uncertainty in the model.

**Conclusions:** Licensing and royalty fees are the primary driver of mRNA-based CGT production costs and represent the greatest opportunity for cost reduction through policy intervention. Strategic priorities for cost reduction should focus on optimizing reagent utilization, increasing platform potency, and expanding use of contract development and manufacturing organizations (CDMOs) to reduce capital and labor costs.

**Key Points:** Producing an example mRNA gene therapy costs about $56 per dose to manufacture, driven almost entirely driven by fees paid to patent holders for the underlying technology. Licensing and royalty fees cost roughly 83 cents of every dollar spent on these new biopharmaceutical products. Until that changes, the gap between what therapies cost to make and what patients and payers are charged will remain very wide.

## 1 Introduction

Cell and gene therapies (CGT) have emerged as a clinically significant modality for conditions ranging from inherited genetic disorders to cancer, offering the potential for durable or curative outcomes with a single treatment or small number of administrations [1, 2]. The approval of mRNA-based vaccines during the COVID-19 pandemic demonstrated that mRNA platforms could be rapidly developed, scaled, and deployed at a global level [3, 4]. These developments have accelerated interest in therapeutic mRNA applications, including gene therapies targeting rare diseases and oncology indications [5, 6].

Despite the clinical promise of these therapies, the high cost of production remains a critical barrier to patient access and commercial viability. Several approved CGT products carry list prices exceeding $1 million per treatment, raising substantial concerns about affordability, reimbursement, and health system sustainability [7, 8]. While a portion of CGT pricing reflects development costs and risk-adjusted return on investment, manufacturing costs, the cost of goods sold (COGS) represents an important and potentially modifiable component of final therapy prices [9].

COGS analysis has been applied productively in pharmaceutical manufacturing to guide investment decisions, benchmark efficiency, and inform pricing negotiations [10, 11]. Within CGT manufacturing, COGS modeling is still maturing; most published analyses are process-specific or proprietary, and relatively little is available in the public literature on the detailed cost structure of mRNA-based gene therapies at commercial scale [12, 13].

The present study addresses this gap by constructing a transparent, scenario-based COGS model for two mRNA-based CGT products: an mRNA vaccine and a therapeutic mRNA gene therapy. We focus specifically on the commercial scale, using publicly available inputs supplemented by expert elicitation, and we apply sensitivity analysis to identify the cost components with the greatest leverage for cost reduction.

Our analysis has direct implications for policy and for manufacturers seeking to reduce COGS. The relative contributions of licensing, royalty fees, materials, consumables, capital equipment, and labor are quantified under base-case, worst-case, and best-case assumptions. The results highlight a structural feature of mRNA-based CGT manufacturing that has received limited attention in the literature: the dominant role of licensing and royalty fees in overall COGS.

## 2 Methods

### 2.1 Modeling Framework

We applied a cost-of-goods-sold modeling approach, which quantifies the direct costs of producing a finished therapeutic product at manufacturing scale. COGS models disaggregate total production cost into its component inputs, including materials, consumables, labor, capital, and overhead, and are widely used in pharmaceutical and biotechnology manufacturing to support business planning and pricing strategy [10, 11].

Our model was implemented in Microsoft Excel and structured around the major manufacturing steps in mRNA-based CGT production: plasmid linearization, mRNA transcription and capping (in vitro transcription, IVT), formulation and encapsulation (including lipid nanoparticle, LNP, preparation), aseptic fill and finish, refrigeration and storage, and quality control (QC). Cost was tracked at each step across six cost categories: (1) material costs, (2) consumable costs, (3) capital costs, (4) labor costs, (5) licensing fees, and (6) royalty fees. Licensing and royalty fees are treated as distinct categories because they arise from different contractual relationships. Licensing fees cover GMP-grade proprietary inputs (e.g., modified nucleotides, LNP formulations), while royalty fees reflect per-unit or per-revenue payments to intellectual property holders.

### 2.2 Products Modeled

Two mRNA-based products were modeled:

1. **mRNA Vaccine:** A prophylactic mRNA-based vaccine, analogous to licensed COVID-19 mRNA vaccines, using LNP delivery. Per-dose mRNA content ranges from 30 to 100 *µ*g across scenarios.
2. **Therapeutic mRNA Gene Therapy:** A therapeutic mRNA product for a non-vaccine indication, with per-dose mRNA content ranging from 0.1 to 1 mg across scenarios.

The therapeutic mRNA product requires substantially higher per-dose mRNA content than the mRNA vaccine, resulting in meaningfully different material and licensing cost profiles.

### 2.3 Production Scale and Facility Assumptions

All modeling was conducted at commercial scale, defined as batch sizes of 10–1,000 liters, with a nominal batch size of 15 liters. The model assumes a large facility (*≥*200 full-time equivalents) operating continuous manufacturing (as opposed to batch manufacturing), which introduces process efficiencies relative to smaller facilities. We assumed 43 batches per production run and 240 working days per year, with a managerial ratio of 1 supervisor per 8 direct-labor employees. GMP-grade materials were assumed to cost five times the equivalent research-grade price when direct vendor quotes were unavailable, consistent with industry conventions [14].

### 2.4 Data Sources and Inputs

Cost inputs were assembled from three primary sources:

1. **Vendor pricing:** Direct vendor quotes and publicly available catalog pricing for GMP-grade materials, consumables, and specialized enzymes.
2. **Published literature:** Peer-reviewed articles and grey literature reporting manufacturing costs, material usage rates, and process yields for mRNA-based products [3, 12, 13].
3. **Subject matter expert consultation:** Input from CGT manufacturing specialists on process assumptions, licensing structures, and industry norms.

Licensing costs were modeled at the component level, including specialized enzymes, modified nucleotides, lipid nanoparticles, cap analogs, standard buffers and salts, cryoprotectants, and plasmid DNA production. Royalty fees were modeled as a percentage of endproduct revenue, with sales per batch assumed at $60,000,000 under base-case assumptions.

### 2.5 Scenario Analysis

Three scenarios were constructed to capture parameter uncertainty:

- **Worst case:** High material costs, high licensing and royalty rates, lower process efficiency, and higher dosage requirements.
- **Base case:** Central estimates for all inputs, reflecting expected commercial performance.
- **Best case:** Lower material costs, favorable licensing terms, higher process efficiency, and lower dosage requirements.

Sensitivity analysis was conducted using a tornado diagram, in which each cost category’s range (from best to worst case) is displayed in descending order of absolute impact on total COGS. This approach identifies which parameters most influence cost outcomes and therefore represent the highest-leverage targets for cost reduction.

### 2.6 Outcome Measures

The primary outcome was total COGS per production run (in US dollars), disaggregated by cost category and manufacturing step. Secondary outcomes included cost per dose and each cost category’s percentage share of total COGS. These metrics were calculated across all three scenarios.

## 3 Results

### 3.1 Total Cost of Goods Sold

Total COGS under base-case assumptions was $16,825,597 per production run. This com-pares to $28,741,463 under worst-case assumptions and $8,289,013 under best-case assumptions, which represents a range of more than 3.5-fold between worst and best cases, which reflects the high sensitivity of mRNA-based CGT manufacturing costs to key input assumptions.

### 3.2 Cost per Dose

Under base-case assumptions, the estimated cost per dose was $56.09. This ranged from $3.68 under best-case assumptions to $383.22 under worst-case assumptions (Table 1). The large range, which is more than 100-fold between best and worst cases, primarily reflects variation in licensing and royalty fee structures, as described below.

**Table 1:** Estimated cost per dose by scenario and cost category (therapeutic mRNA, commercial scale, 15 L batch, 43 batches)

| Cost Category | Worst Case (\$) | Base Case (\$) | Best Case (\$) |
| --- | --- | --- | --- |
| Consumable Costs | 14.83 | 3.26 | 0.42 |
| Material Costs | 26.38 | 5.80 | 0.74 |
| Capital Costs | 0.55 | 0.11 | 0.01 |
| Labor Costs | 2.25 | 0.39 | 0.03 |
| License Fees | 185.60 | 23.32 | 1.41 |
| Royalty Fees | 153.60 | 23.20 | 1.08 |
| **Total** | **383.22** | **56.09** | **3.68** |

### 3.3 Cost Breakdown by Category

Table 2 and Figure 1 present total COGS per production run by cost category across all three scenarios. Under base-case assumptions, licensing fees and royalty fees together account for $13,956,000 of $16,825,597 in total COGS, or approximately 82.9% of total costs. License fees alone represent 41.6% of total COGS ($6,996,000), and royalty fees represent an additional 41.4% ($6,960,000).

**Figure 1:**
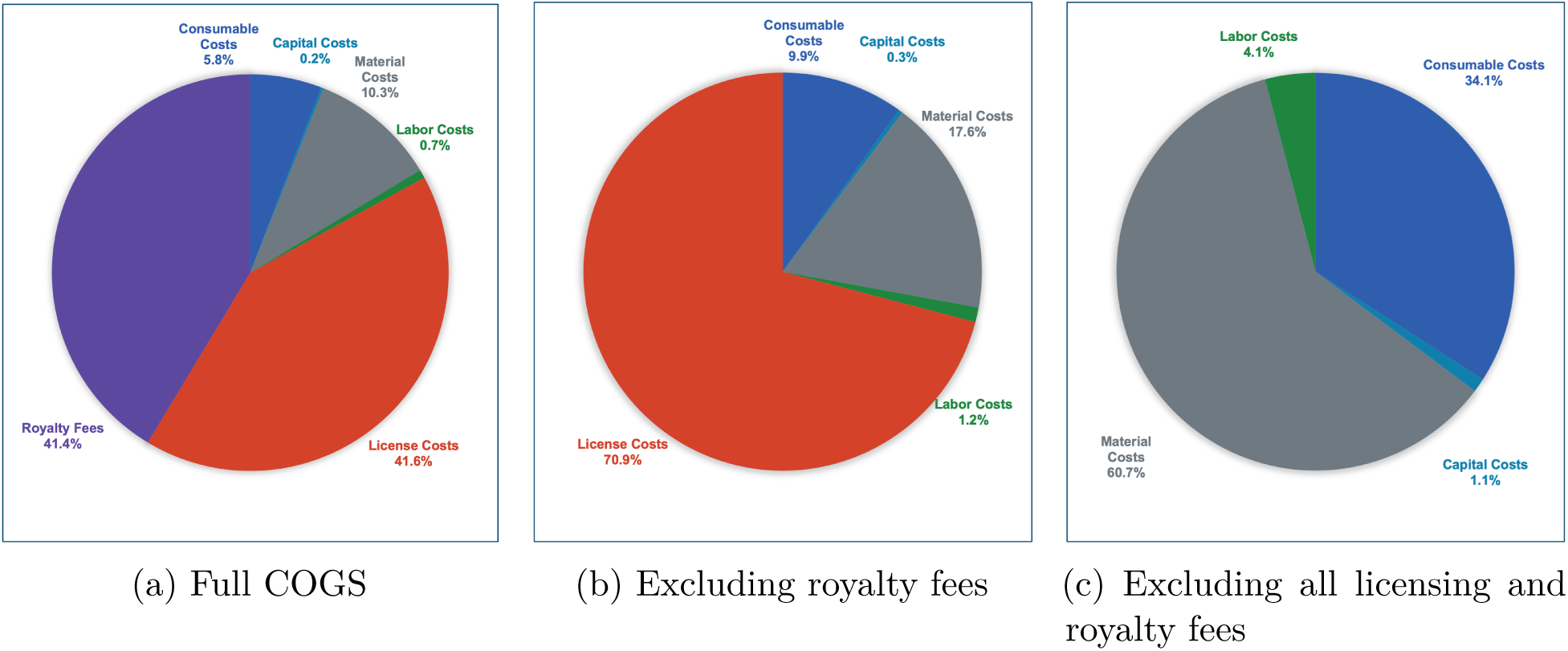
Base-case COGS by cost category for a commercial-scale therapeutic mRNA gene therapy (15 L batch, 43 batches). Panel (a) shows full COGS; license and royalty fees together account for 82.9% of total costs ($16,825,597). Panel (b) removes royalty fees, leaving license costs dominant at 70.9%. Panel (c) removes all fees, revealing that materials (60.7%) and consumables (34.1%) are the largest controllable cost drivers.

**Table 2:** Total COGS per production run by cost category and scenario.

| Cost Category | Worst Case |  | Base Case |  | Best Case |  |
| --- | --- | --- | --- | --- | --- | --- |
| | \$ | % | \$ | % | \$ | % |
| Consumables | 1,112,470 | 3.9 | 978,976 | 5.8 | 934,475 | 11.3 |
| Materials | 1,978,579 | 6.9 | 1,741,149 | 10.3 | 1,662,006 | 20.1 |
| Capital | 41,523 | 0.1 | 32,188 | 0.2 | 26,172 | 0.3 |
| Labor | 168,891 | 0.6 | 117,284 | 0.7 | 68,360 | 0.8 |
| License | 13,920,000 | 48.4 | 6,996,000 | 41.6 | 3,168,000 | 38.2 |
| Royalty | 11,520,000 | 40.1 | 6,960,000 | 41.4 | 2,430,000 | 29.3 |
| **Total** | **28,741,463** | **100** | **16,825,597** | **100** | **8,289,013** | **100** |

**Figure 2:**
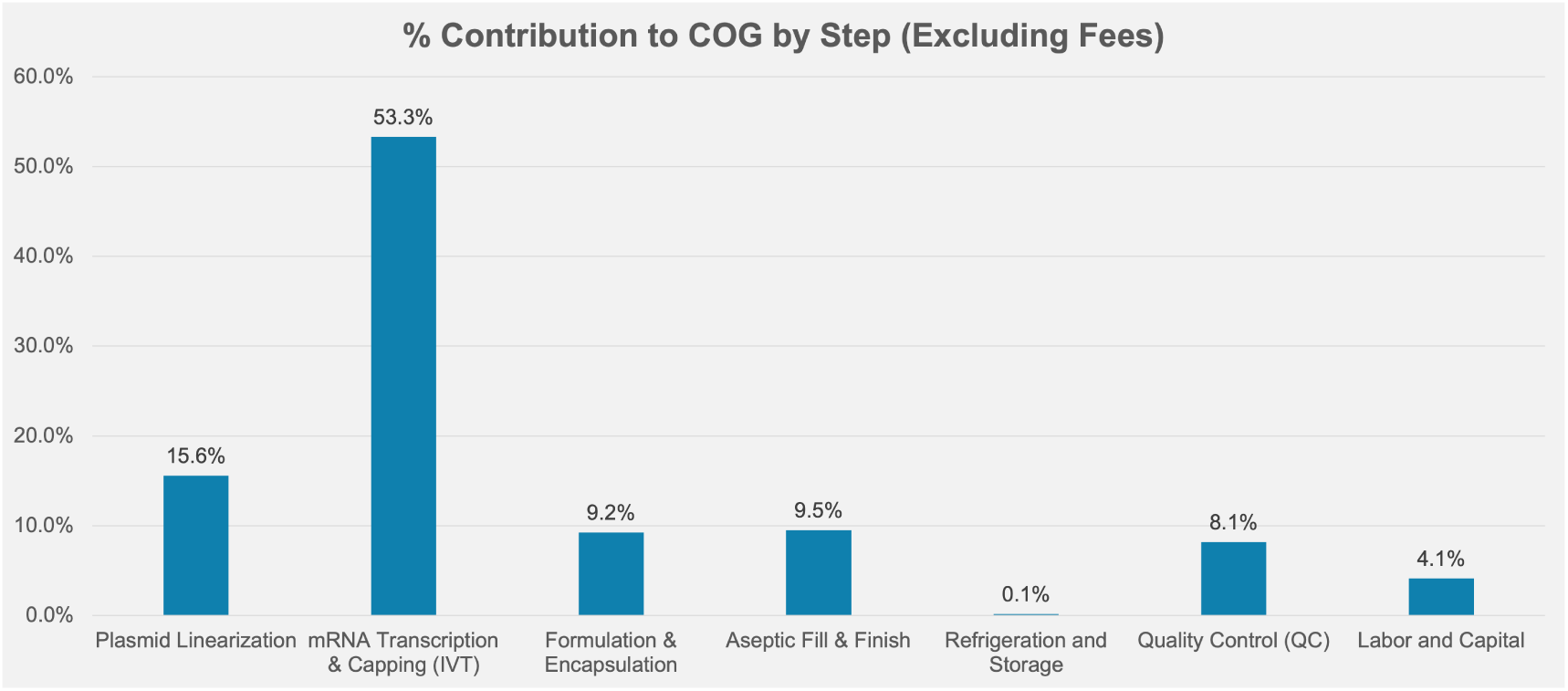
Percentage contribution to COGS by manufacturing step, excluding licensing and royalty fees. mRNA transcription and capping (IVT) accounts for 53.3% of non-licensing COGS, followed by aseptic fill and finish (9.5%), plasmid linearization (15.6%), formulation and encapsulation (9.2%), quality control (8.1%), and labor and capital (4.1%).

Excluding licensing and royalty fees, the remaining production costs are driven by material costs (61% of non-licensing COGS, or $1,741,149), consumable costs (34%, or $978,976), capital costs (4%, or $32,188), and labor costs (1%, or $117,284).

### 3.4 Cost Breakdown by Manufacturing Step

Table 3 shows COGS by manufacturing step. The largest process-level cost is incurred at the mRNA transcription and capping (IVT) step, which accounts for 8.6% of base-case total COGS ($1,439,102). Plasmid linearization (3.2%), formulation and encapsulation (1.6%), and aseptic fill and finish (1.6%) represent the next largest steps. The majority of COGS under base-case assumptionsis attributed to “paid at end” licensing and royalty costs that are assessed across the production run rather than at a specific manufacturing step (83%).

**Table 3:**
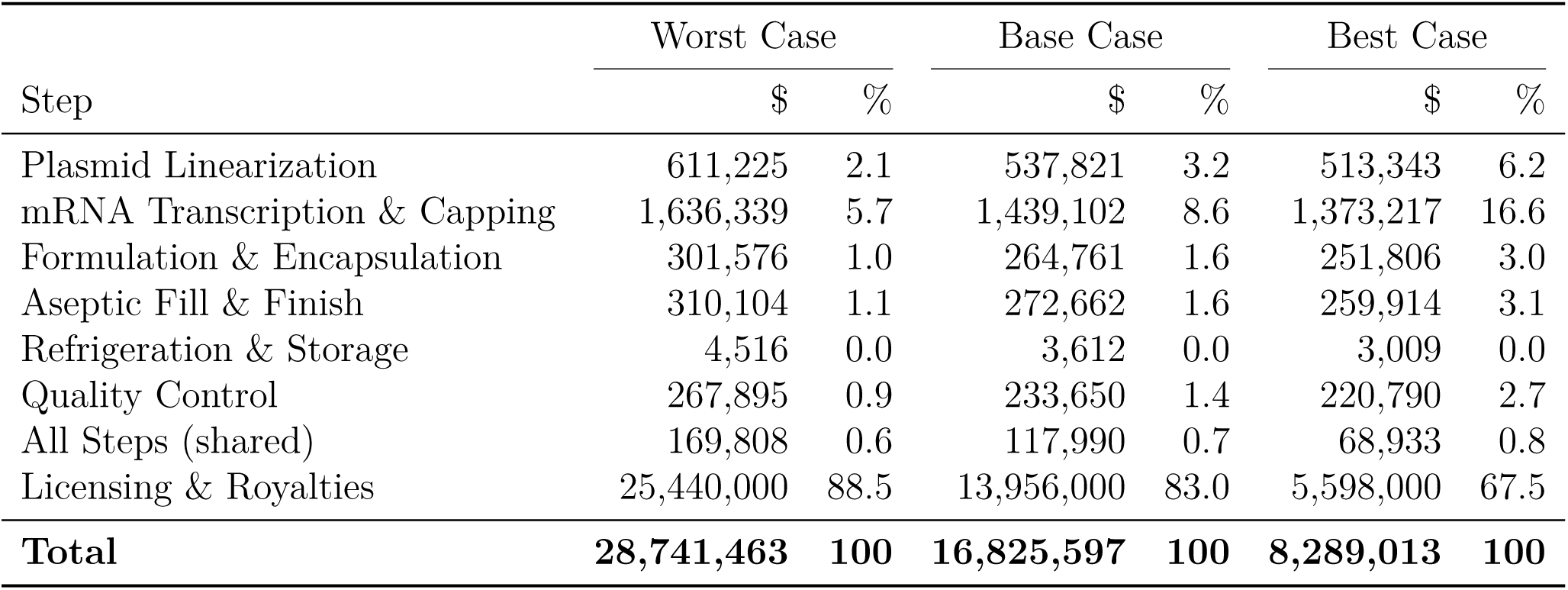
Total COGS per production run by manufacturing step and scenario.

| Step | Worst Case |  | Base Case |  | Best Case |  |
| --- | --- | --- | --- | --- | --- | --- |
| | \$ | % | \$ | % | \$ | % |
| Plasmid Linearization | 611,225 | 2.1 | 537,821 | 3.2 | 513,343 | 6.2 |
| mRNA Transcription & Capping | 1,636,339 | 5.7 | 1,439,102 | 8.6 | 1,373,217 | 16.6 |
| Formulation & Encapsulation | 301,576 | 1.0 | 264,761 | 1.6 | 251,806 | 3.0 |
| Aseptic Fill & Finish | 310,104 | 1.1 | 272,662 | 1.6 | 259,914 | 3.1 |
| Refrigeration & Storage | 4,516 | 0.0 | 3,612 | 0.0 | 3,009 | 0.0 |
| Quality Control | 267,895 | 0.9 | 233,650 | 1.4 | 220,790 | 2.7 |
| All Steps (shared) | 169,808 | 0.6 | 117,990 | 0.7 | 68,933 | 0.8 |
| Licensing & Royalties | 25,440,000 | 88.5 | 13,956,000 | 83.0 | 5,598,000 | 67.5 |
| **Total** | **28,741,463** | **100** | **16,825,597** | **100** | **8,289,013** | **100** |

### 3.5 Licensing Cost Components

Within total licensing costs, lipid nanoparticle (LNP) formulation licenses represent by far the largest single component. Under base-case assumptions, LNP licensing alone accounts for $6,000,000 per production run out of $6,996,000 in total license fees (approximately 86%), at a per-batch sales assumption of $60,000,000. Specialized enzymes ($300,000) and modified nucleotides ($300,000) each contribute substantially smaller shares. Standard buffers, salts, and dNTPs ($18,000), cap analogs ($18,000), cryoprotectants and stabilizers ($330,000), and plasmid DNA production licenses ($30,000) round out the license cost structure.

Licensing costs scale with product revenue rather than with production volume alone, because they are structured as a percentage of sales. This creates a situation in which high-value therapies face disproportionately high licensing burdens, and cost-per-dose reduction through volume scale-up provides limited relief.

### 3.6 Sensitivity Analysis

Figure 3 confirms that licensing and royalty fees exhibit by far the largest sensitivity range of any cost category. The range from best to worst case for licensing alone spans approximately $10,752,000 per production run, and for royalty fees spans approximately $9,090,000. Combined, these two categories drive a COGS range of approximately $19,842,000 between best and worst case compared to roughly $630,000 for all other cost categories combined.

**Figure 3:**
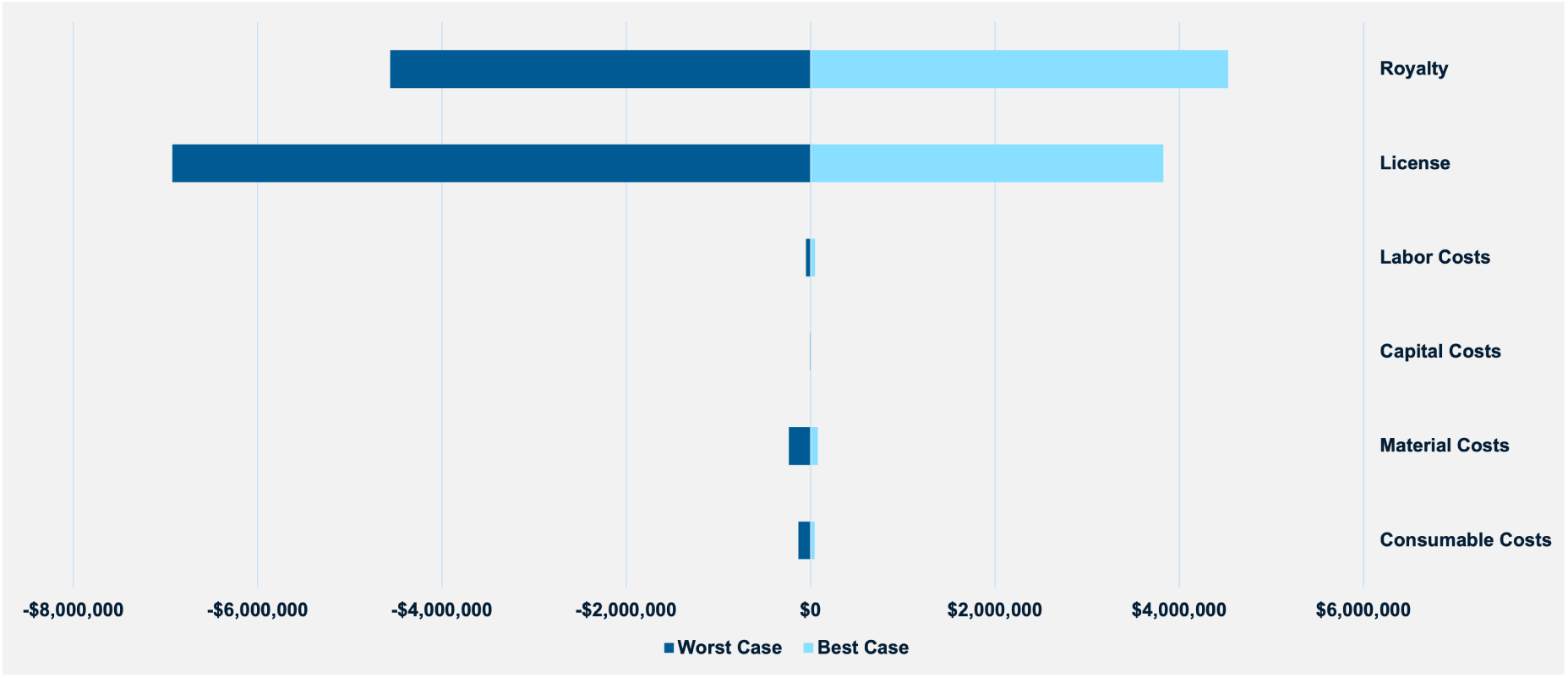
Tornado diagram showing the change in total COGS from the base case under worst-case (red, leftward) and best-case (blue, rightward) assumptions for each cost category. License fees and royalty fees together account for over 97% of total COGS uncertainty across scenarios; all other cost categories are negligible by comparison on this scale.

**Figure 4:**
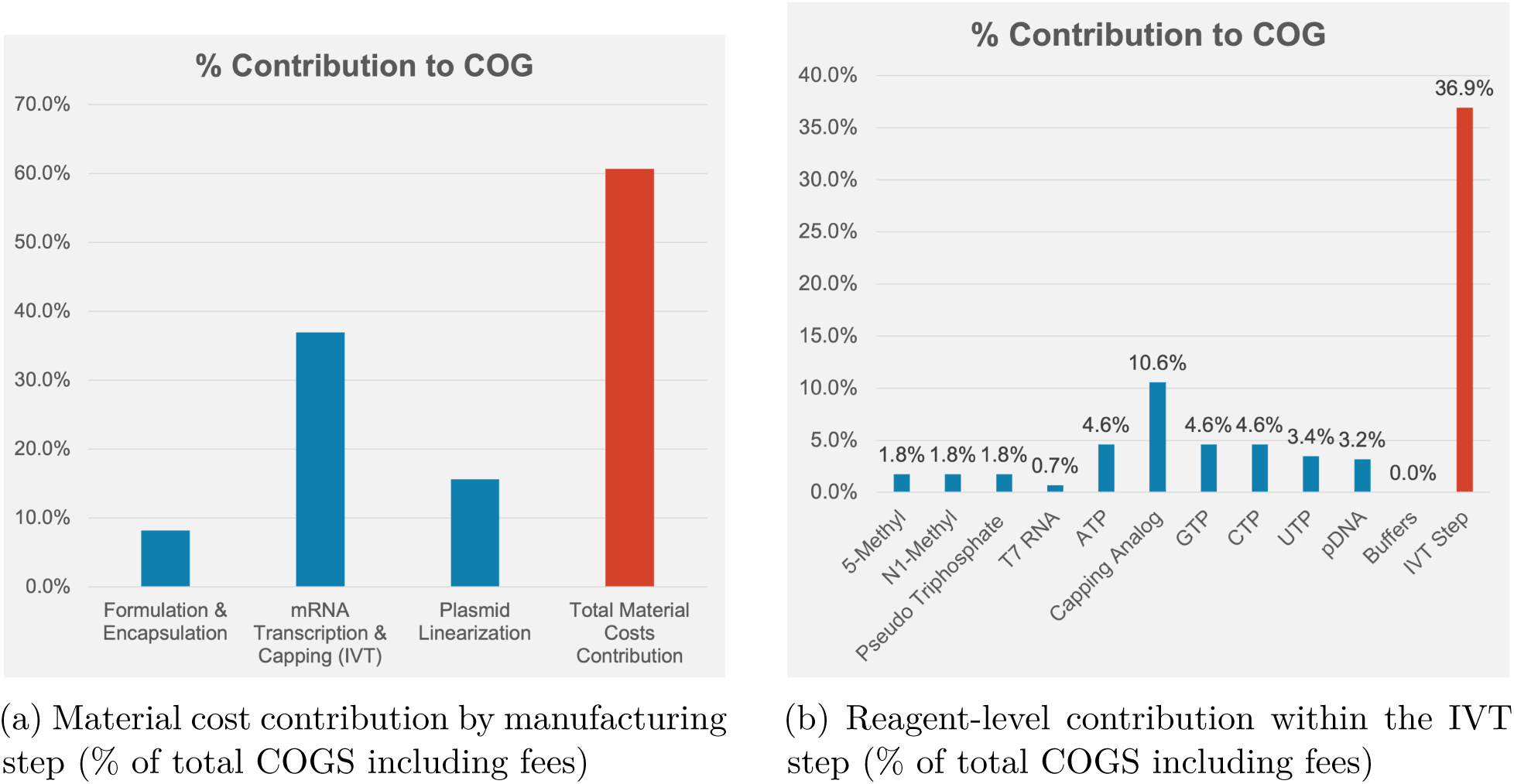
Material cost drivers under base-case assumptions. Panel (a): mRNA transcription and capping (IVT) accounts for 36.9% of total COGS, representing by far the most material-intensive step. Panel (b): within IVT, capping analog (10.6%) is the single largest reagent cost, followed by ATP, GTP, and CTP (4.6% each) and UTP (3.4%).

Among non-licensing cost categories, material costs show the widest sensitivity range, followed by consumables, labor, and capital (see Supplementary Figure S1 for the tornado diagram restricted to non-licensing categories). Capital and labor costs contribute minimally to total COGS uncertainty at commercial scale, reflecting the efficiency gains associated with large continuous manufacturing facilities and the shift to CDMOs.

## 4 Discussion

### 4.1 Licensing and Royalty Fees as the Dominant Cost Driver

The central finding of this analysis is that licensing and royalty fees account for the overwhelming majority of COGS for mRNA-based CGT at commercial scale, reaching approximately 83% of total costs under base-case assumptions. This finding is consistent with the structure of intellectual property in the mRNA and LNP delivery space, where a small number of organizations hold foundational patents governing modified nucleotide synthesis, LNP formulation, and IVT reaction chemistry [15, 16].

The dominant role of LNP licensing is particularly notable. Lipid nanoparticle delivery systems are essential for in vivo delivery of mRNA, and the patent landscape in this space remains concentrated [17]. Licensing fees for LNP technology can be structured as a percentage of net revenue or as a fixed per-dose fee, and the former approach which was modeled here, creates a direct link between product price and licensing cost. This structure has the consequence that reducing per-dose production costs does not proportionally reduce per-dose licensing costs, limiting the leverage that manufacturing optimization provides.

Royalty fees compound this challenge. Unlike license fees for GMP-grade inputs, royalty fees reflect ongoing payments to intellectual property holders based on product commercialization. Under the base-case assumptions modeled here, royalty fees are approximately equal in magnitude to license fees ($6,960,000 vs. $6,996,000). Together, they represent a structural cost floor that cannot be addressed through manufacturing efficiency improvements alone.

### 4.2 Implications for Non-Licensing Costs

Among the costs that manufacturers can more directly influence, material costs represent the largest lever (61% of non-licensing COGS). The mRNA transcription and capping step, which is the most material-intensive step in the production process, is also the highest-cost manufacturing step. Key input materials at this step include modified nucleotides (e.g., N1-methylpseudouridine), IVT enzymes, and plasmid DNA template. Strategies for reducing material costs at IVT include improving nucleotide incorporation efficiency, reducing batch-level material wastage, and negotiating volume discounts for GMP-grade inputs.

Consumable costs (34% of non-licensing COGS) represent the second-largest category. These include single-use bioreactor components, filtration assemblies, and disposable process equipment. The shift to single-use systems has been broadly adopted in CGT manufacturing for biosafety and flexibility reasons, but this comes with ongoing consumable expenditure that can be meaningfully reduced through supply chain consolidation and platform standardization.

Capital and labor costs together represent approximately 5% of non-licensing COGS under base-case assumptions. This finding reflects the economies of scale available at large commercial facilities (*≥*200 FTEs) operating continuous manufacturing processes. The expanded use of CDMOs has been particularly effective in reducing capital costs: manufacturers outsourcing to CDMOs avoid the fixed capital investment in dedicated facilities and can access existing GMP-certified production capacity [18]. Labor costs are further reduced at large continuous facilities through automation and process integration.

### 4.3 Cost-per-Dose Considerations

The estimated base-case cost per dose of $56.09 must be interpreted in context. This figure represents the cost to manufacture one dose of therapeutic mRNA at commercial scale, and it does not include development costs, clinical trial expenses, regulatory costs, distribution, or profit margin. List prices for approved mRNA-based gene therapies have substantially exceeded manufacturing costs, reflecting these additional elements [7]. Nonetheless, identifying the manufacturing cost floor is important for policy analysis, because it establishes the lower bound below which prices could not fall without losses on production alone.

The 100-fold range between best-case ($3.68) and worst-case ($383.22) cost-per-dose estimates demonstrates the degree of uncertainty that currently characterizes this space. Much of this uncertainty reflects the variability of licensing and royalty terms across developers, which are often proprietary and not disclosed. Public policy interventions that increase transparency in CGT licensing markets, analogous to efforts to increase transparency in pharmaceutical supply chain pricing more broadly, could reduce this uncertainty and enable more accurate cost benchmarking.

### 4.4 Policy Implications

Three policy implications follow from these results. First, because licensing and royalty fees represent the largest share of mRNA-based CGT production costs, government action to support licensing reform or create shared intellectual property pools could have a more substantial impact on manufacturing costs than investments in manufacturing efficiency alone.

Second, strategic investment in platform potency, increasing the therapeutic effect per unit of mRNA dosed, can reduce total material requirements and thereby lower the mRNA content required per dose. This approach targets both direct material costs and the dosage-sensitive component of licensing fees. Improvements in LNP delivery efficiency, for example, could reduce effective dose requirements and lower both material and licensing costs simultaneously.

Third, expanded use of CDMOs has demonstrably reduced capital and labor costs for CGT manufacturers. Policy mechanisms that support the development of CDMO capacity for advanced therapy manufacturing, including workforce development, facility investment credits, and regulatory harmonization for CDMO-client technology transfer, can reinforce this trend.

### 4.5 Limitations

Several limitations should be noted. First, the model focuses on a single production scenario (15 L batch, 43 batches, large continuous facility) and may not generalize to smaller-scale or differently structured production operations. Second, licensing and royalty cost inputs are drawn from published sources and expert consultation, and actual proprietary licensing terms may differ substantially from modeled values. The high sensitivity of results to these inputs as demonstrated by the tornado analysis underscores the importance of this limitation. Third, the model does not incorporate dynamic considerations such as yield improvements over time, changes in input prices, or the effects of competition on licensing terms. Fourth, the two products modeled (mRNA vaccine and therapeutic mRNA) represent a subset of the CGT landscape; cell therapies, viral vector gene therapies, and other modalities would exhibit different cost structures.

## 5 Conclusions

This analysis provides a transparent, scenario-based estimate of COGS for mRNA-based CGT at commercial scale, with specific attention to the structure of licensing and royalty costs. Under base-case assumptions, licensing and royalty fees account for approximately 83% of total COGS, dwarfing all other cost categories. Among non-licensing costs, materials and consumables are the primary drivers. Cost per dose is estimated at $56.09 under base-case assumptions, with a range from $3.68 to $383.22 across scenarios.

These findings suggest that strategies to reduce CGT manufacturing costs must address licensing and royalty structures, which are not amenable to efficiency-based optimization. Policy interventions focused on licensing reform, shared IP frameworks, and increased pricing transparency have the potential to reduce the structural cost floor for mRNA-based therapies. Simultaneously, manufacturers should continue to invest in platform potency and CDMO partnerships, which offer meaningful leverage over the non-licensing share of COGS.

Future work should extend this modeling approach to other CGT modalities, incorporate dynamic cost trajectories, and examine the relationship between manufacturing COGS and final therapy pricing in greater detail.

## Statements and Declarations

### Funding

This work was supported by a contract from the Advanced Research Projects Agency for Health (ARPA-H) to The MITRE Corporation.

## Competing Interests

The authors declare no competing interests.

## Ethics Approval

Not applicable. This study did not involve human participants, human data, or biological material.

## Use of Artificial Intelligence

AI-assisted copy editing was used in the preparation of this manuscript.

## Author Contributions

R.D.L. conceived and led the analysis, built the cost model, and drafted the manuscript. P.B., B.H., A.O., M.W., and A.W. contributed to model design, data collection, and critical revision of the manuscript. All authors reviewed and approved the final version.

## Data Availability

The Excel-based COGS model supporting this analysis is available from the corresponding author upon reasonable request. The mRNA vaccine model results are alos available from the corresponding author upon request.

## Prior Presenation

Preliminary model results were presented at the American Public Health Association Annual Meeting in Washington, DC in November, 2025.

## Supplementary Material

**Figure 5:**
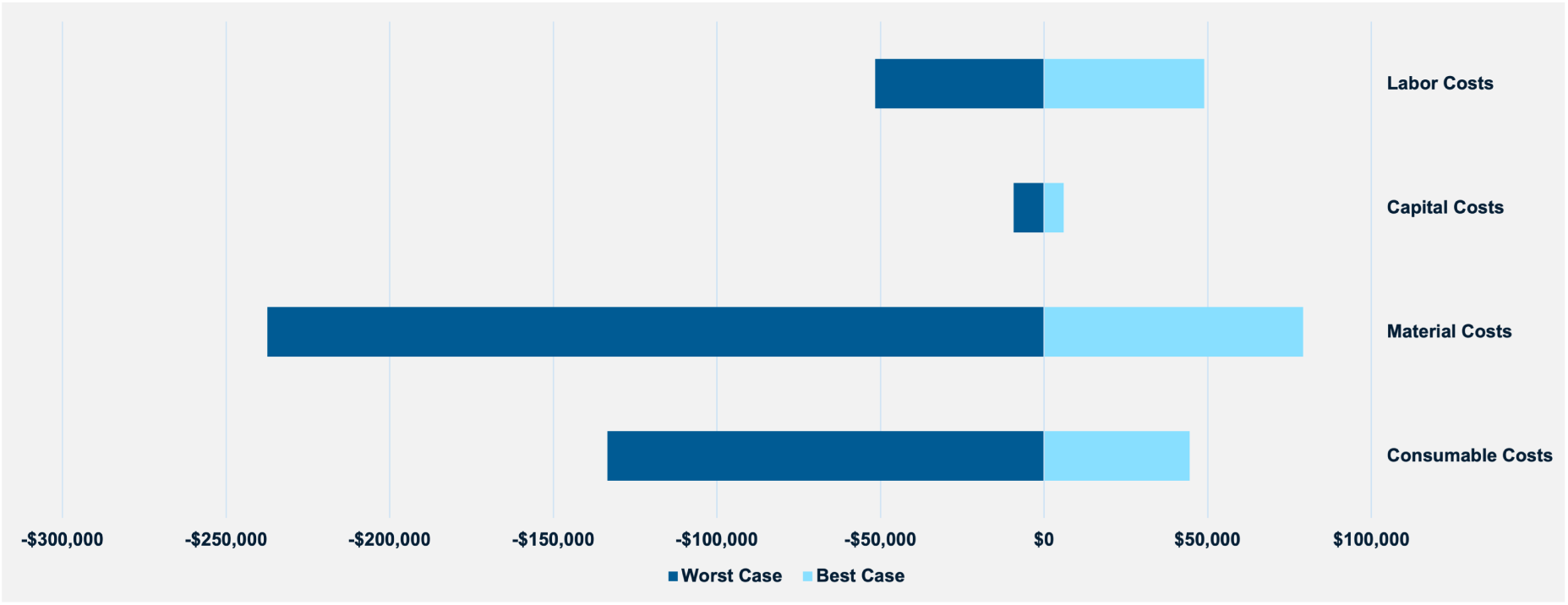
**Supplementary Figure S1.** Tornado diagram for non-licensing cost categories only (materials, consumables, labor, and capital), showing the change in COGS from the base case under worst-case (red) and best-case (blue) assumptions. Material costs ($237K worst-case deviation) represent the largest non-licensing lever, followed by consumables ($133K) and labor ($52K). Capital costs ($9K range) are negligible at commercial scale. The combined worst-to-best range for all four categories is approximately $630,000, compared to $19.8 million for licensing and royalty fees combined (Figure 3).

## Data Availability

The minimal data set and Excel spreadsheet is available from the author upon request.

## NOTICE

This software/technical data was produced for the U.S. Government under Contract Number 75FCMC18D0047/75FCMC23D0004, and is subject to Federal Acquisition Regulation Clause 52.227-14, Rights in Data-General.

No other use other than that granted to the U.S. Government, or to those acting on behalf of the U.S. Government under that Clause is authorized without the express written permission of The MITRE Corporation.

For further information, please contact The MITRE Corporation, Contracts Management Office, 7515 Colshire Drive, McLean, VA 22102-7539, (703) 983-6000.

## Notes

### Competing Interest Statement

The authors have declared no competing interest.

### Funding Statement

This study was funded by the Advanced Research Projects Agency for Health, ARPA-H.

### Summary of Updates

Copyright notice added to comply with institutional requirements.

